# Genome wide association study of human bacteremia *Escherichia coli* isolates identifies genetic determinants for the portal of entry but not fatal outcome

**DOI:** 10.1101/2021.11.09.21266136

**Authors:** Erick Denamur, Bénédicte Condamine, Marina Esposito-Farèse, Guilhem Royer, Olivier Clermont, Cédric Laouenan, Agnès Lefort, Victoire de Lastours, Marco Galardini, the COLIBAFI, SEPTICOLI groups

## Abstract

*Escherichia coli* is an important cause of bloodstream infections (BSI), which is of concern given its high mortality and increasing worldwide prevalence. Finding bacterial genetic variants that might contribute to patient death is of interest to better understand its mechanism and implement diagnostic methods that specifically look for those factors. *E. coli* samples isolated from patients with BSI are an ideal dataset to systematically search for those variants, as long as the influence of host factors such as comorbidities are taken into account. Here we performed a genome-wide association study (GWAS) using data from 910 patients with *E. coli* BSI from hospitals in Paris, France; we looked for associations between bacterial genetic variants and three patient outcomes (death at 28 days, septic shock and admission to intensive care unit), as well as two portals of entry (urinary and digestive tract), using various clinical variables from each patient to account for host factors. We did not find any associations between genetic variants and patient outcomes, potentially confirming the strong influence of host factors in influencing the course of BSI; we however found a strong association between the *papGII*/*papGIII* operon and entrance of *E. coli* through the urinary tract, which demonstrates the power of bacterial GWAS even when applied to actual clinical data. Despite the lack of associations between *E. coli* genetic variants and patient outcomes, we estimate that increasing the sample size by one order of magnitude could lead to the discovery of some putative causal variants. The adoption of bacterial genome sequencing of clinical isolates might eventually lead to the elucidation of the mechanisms behind BSI progression and the development of sequence-based diagnostics.

## Introduction

*Escherichia coli* bloodstream infections (BSI) represent an increasing public health burden as (i) they exhibit high mortality (between 10 and 30%)^1,2^, (ii) its worldwide prevalence is increasing since the 2000s^3^ and (iii) antimicrobial resistance is rising in *E. coli*^*3*^, which could impact patients’ management and infection outcome. Molecular epidemiology of BSI has been refined in the last few years thanks to whole genome sequencing. *E. coli* has a clonal population structure^4^ with the delineation of at least eight phylogroups (A, B1, B2, C, D, E, F and G)^5^. Strains responsible for BSI belong mainly to a few clonal lineages including sequence type (ST) 131, ST73 and ST95, all of the B2 phylogroup, ST69 (D phylogroup) and ST10 (A phylogroup)^5^. Until now, classical multivariate analyses have identified host factors and portal of entry as the major determinants of a patient’s death, while bacterial genetic traits have been associated with a smaller effect size to mortality or only in a subset of studies^1,2,6–10^.

Bacterial genome wide association studies (GWAS) are now common thanks to an increase in sequencing capacity and specific computational tools^11^; in *E. coli* they have allowed the identification of genetic traits linked to pathogenicity in avian strains^12^, invasiveness in urinary tract infection (UTI) strains^13^ and isolation source^14^. However, they failed to identify genetic markers of disease severity in Shigellosis^15^. This could have multiple explanations: disease severity (e.g. patient death) is a trait that is not under selection as it doesn’t provide a reproductive advantage, and is therefore less likely to evolve independently across multiple lineages, which in turn makes it less likely to be found through bacterial GWAS^16^. Furthermore, as opposed to antimicrobial resistance which is often caused by a handful of genetic variants, disease severity might involve multiple genetic loci, each with small effects, which are harder to discover. Lastly, small sample sizes can lead to insufficient power to find causal variants.

Identifying microbial genetic elements that contribute to the outcome of BSI is of interest to i) better understand the molecular mechanisms of microbial infection, and ii) improve patient care and prediction of clinically-relevant bacterial traits based on microbial genomics data, which is increasingly becoming available with very low turnaround time^17^. In this context, we performed GWAS on data from two large clinical observational prospective multicentric studies from the Paris area (Septicoli^10^ and Colibafi^8^) involving a total of 910 adult patients with *E. coli* BSI. We used the clinical information from each patient, such as age, comorbidities and treatment as covariates to reduce the influence of host factors in the association analysis^18^. We found no association between microbial genetic elements and infection outcomes, which seems to indicate that host and clinical factors have a more predominant role. On the other hand, we found a clear signal between several genes such as the *pap* operon with the urinary and digestive portals of entry. Lastly, we ran a statistical power analysis using a set of simulated genomes, which indicated how a 10-fold increase in sample size may lead to the discovery of further bacterial factors associated with the establishment of BSI and with its outcomes.

## Results

### A combined dataset of 910 BSI patients with matching clinical data and bacterial isolates whole genomes

In this study we combined data from two similar clinical studies (Colibafi^8^ and Septicoli^10^), conducted across 11 teaching hospitals, belonging to the same institution, the “Assistance Publique-Hôpitaux de Paris” (AP-HP), across and around Paris, France. The earlier study (Colibafi, 2005) originally included 1,051 patients across the whole of France, with information about bacterial genetic determinants obtained through PCR molecular assays; in this study we kept only those 365 samples originating from 8 hospitals from the Paris area to avoid geographical biases. From the later study (Septicoli, 2016-7) we kept all the 545 samples from 7 hospitals in the Paris area. Bacterial genomes of these samples from both studies, generated by Illumina technology, were available^19^. We focused on three outcomes for the patients represented in the combined dataset, namely death at 28th days, presence of a septic shock and admission to an intensive care unit (ICU); we note that these outcomes are not mutually exclusive. We found that the prevalence of these outcomes in the two studies was 10.7%, 24% and 14.5%, for death, septic shock and admission to ICU, respectively (Figure 1a). We found that the prevalence of death and admission to ICU was very similar between the two studies, with 12.3% and 9.5% of deaths in the Colibafi and Septicoli studies, respectively, and admission to the ICU reported for 12.3% and 16% of patients. Conversely, we found a much higher incidence of patients experiencing septic shock in the Septicoli cohort: 32.5% of patients versus 11.2% in Colibafi (Figure 1b). These variations may be due to the different hospitals contributing the clinical data between the two studies: indeed, even though both studies are exclusively focused on the AP-HP teaching hospitals in Paris, only 4 out of 11 hospitals are included in both studies^19^. We additionally focused on the reported portal of entry of the BSI, which has been previously found to be predictive of patient outcome; urinary and digestive tract portals of entry were the most prevalent in the combined dataset - 58.2% and 35.7% of patients, respectively. The other reported portals of entry all had a prevalence below 5% (Figure 1a), and we therefore chose to only use the urinary and digestive tract portals of entry for all subsequent analyses. We found that entry through the digestive tract was reported for 41.8% patients in the Septicoli study, compared to 26.6% in the Colibafi study, which again may be due to differences in the hospitals providing the data for both studies (Figure 1b). The age distribution between the two studies is comparable, with median age of the patients being 67 and 69 years in the Colibafi and Septicoli studies, respectively (Figure 1c). To reduce the influence of these differences between the two studies on our analyses, we introduced the study provenance as a covariant in the combined dataset (Supplementary Table 1).

**Figure 1.**
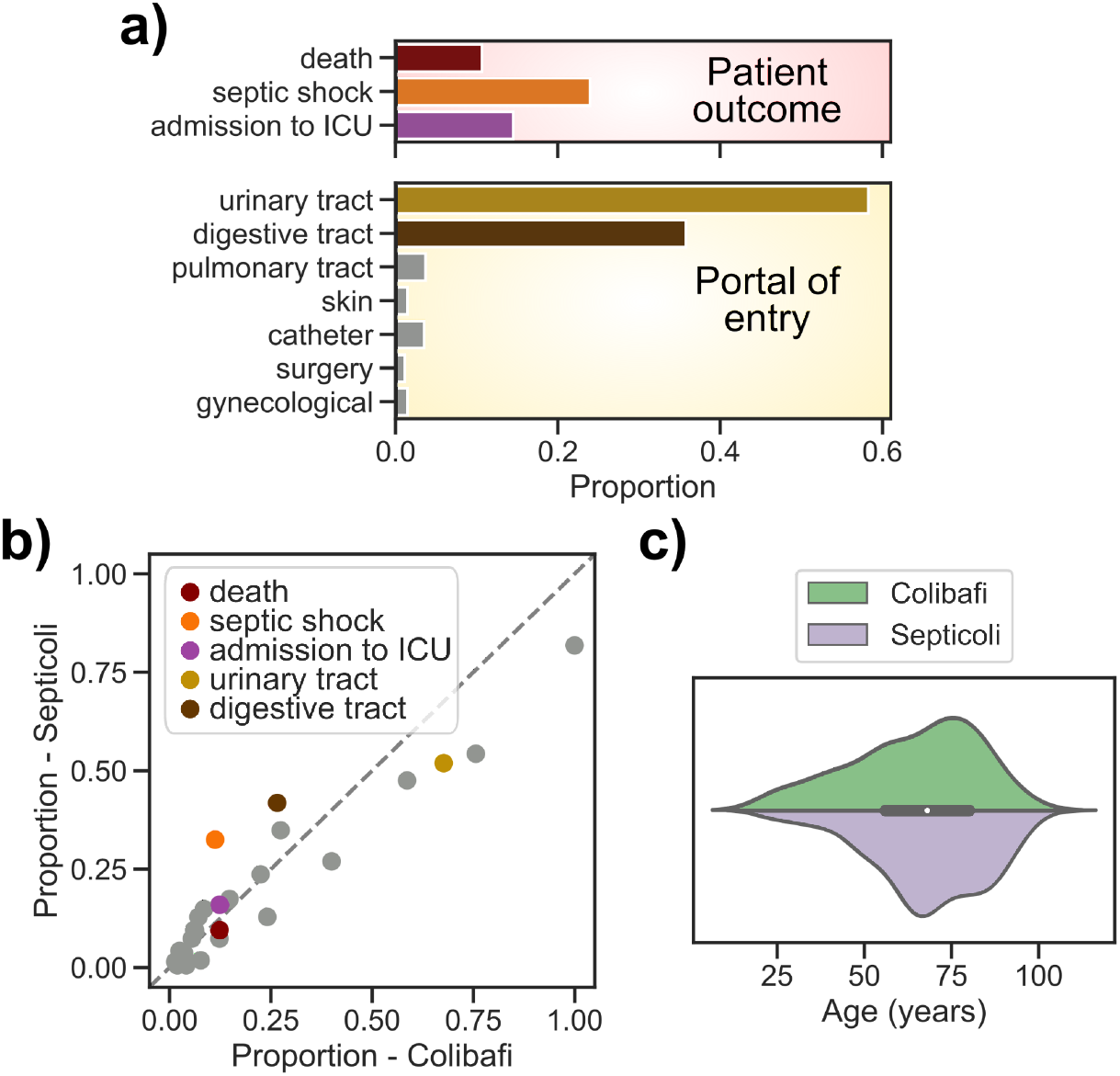
Clinical variables of the combined dataset (910 BSI samples). a) Proportion of the three patient outcomes after BSI and their portals of entry. b) Scatterplot of the proportion of all binary clinical variables in the two studies, highlighting the major differences. c) Violin plot showing the patients’ age distribution between the two studies.

### The pathogen portal of entry is associated with BSI outcomes

We found that several clinical variables are associated with the three patient outcomes, consistent with earlier analyses on the two studies alone^8,10^ (Table 1). Among other variables, we found that entry through the pulmonary and digestive tract were associated with the death of the patient (odds ratio 2.88 and 1.51 and p-values 8E^-5^ and 0.006, respectively), while entry through the urinary tract was found to be protective (odds ratio 0.51, p-value 2E^-5^). We found that entry through the pulmonary tract was also associated with patients experiencing a septic shock (odds ratio 2.12, p-value 4E^-3^), while we found that entry through the digestive tract was associated with patients being admitted to the ICU, among other variables (odds ratio 1.53, p-value 1E^-3^). When combining all clinical variables with association p-value < 0.1 into a multivariate analysis (Table 2, see Methods) we found that portal of entry was again the dominant variable associated with patient outcomes, together with study provenance. In particular we found that entry through the pulmonary tract was significantly associated with a patient’s death (odds ratio 2.40, p-value 0.003), while entry through the urinary tract was negatively associated (odds ratio 0.64, p-value 0.008). We found that entry through the pulmonary tract was also significantly associated with a patient experiencing a septic shock (odds ratio 2.10, p-value 0.005), while entry through the digestive tract was associated with a patient being admitted to the ICU (odds ratio 1.57, p-value 0.002). This analysis underscores the influence of the *E. coli* portal of entry on BSI outcomes, which in turn could have an association with specific bacterial genetic elements.

We found that no *E. coli* phylogroup was associated with patient death (p-value > 0.01), consistent with earlier analyses from the two separate studies^8,10^, in contrast to what we previously observed in a mouse model of BSI, in which we found that the B2 phylogroup was associated with the death of the animal^20^. This difference suggests that host and clinical factors may have a larger influence on patient outcomes than the genetic background of the bacterial isolate, at least in this dataset. We also observed no association between an isolate’s phylogroup and a septic shock or admission to the ICU. The absence of these genetic background effects does not imply that there are no “locus effects”, meaning that individual genetic variants may still be found to be associated with patients’ outcomes. We found a strong association between the isolates’ phylogroup and the urinary and digestive tract portals of entry; phylogroups A, B1 and B2 were associated with both phenotypes (p-value < 0.01, Supplementary Table 2). Such similarity between the two phenotypes is not surprising given the low prevalence of the other portals of entry, leading to two almost mutually exclusive traits (Figure 2).

**Table 1.** Univariate analysis on the combined dataset. Only clinical variables significantly associated with BSI outcomes are shown. CI, confidence interval.

| Patient outcome | Clinical variable | Odds-ratio [95% CI] | P-value |
| --- | --- | --- | --- |
| death | urinary tract | 0.51 [0.38-0.69] | 2E<sup>-5</sup> |
|  | pulmonary tract | 2.88 [1.70-4.87] | 8E<sup>-5</sup> |
|  | malignant tumor | 1.75 [1.30-2.35] | 2E<sup>-4</sup> |
|  | digestive tract | 1.51 [1.12-2.04] | 0.006 |
|  | chronic alcoholism | 1.74 [1.16-2.60] | 0.007 |
|  | immunosuppression | 1.50 [1.11-2.02] | 0.007 |
|  | active smoking | 1.56 [1.11-2.19] | 0.009 |
| septic shock | pulmonary tract | 2.12 [1.27-3.55] | 0.003 |
| admission to ICU | cirrhosis | 1.99 [1.37-2.89] | 2E<sup>-4</sup> |
|  | digestive tract | 1.53 [1.18-1.99] | 0.001 |
|  | active smoking | 1.59 [1.17-2.16] | 0.003 |

**Table 2.**
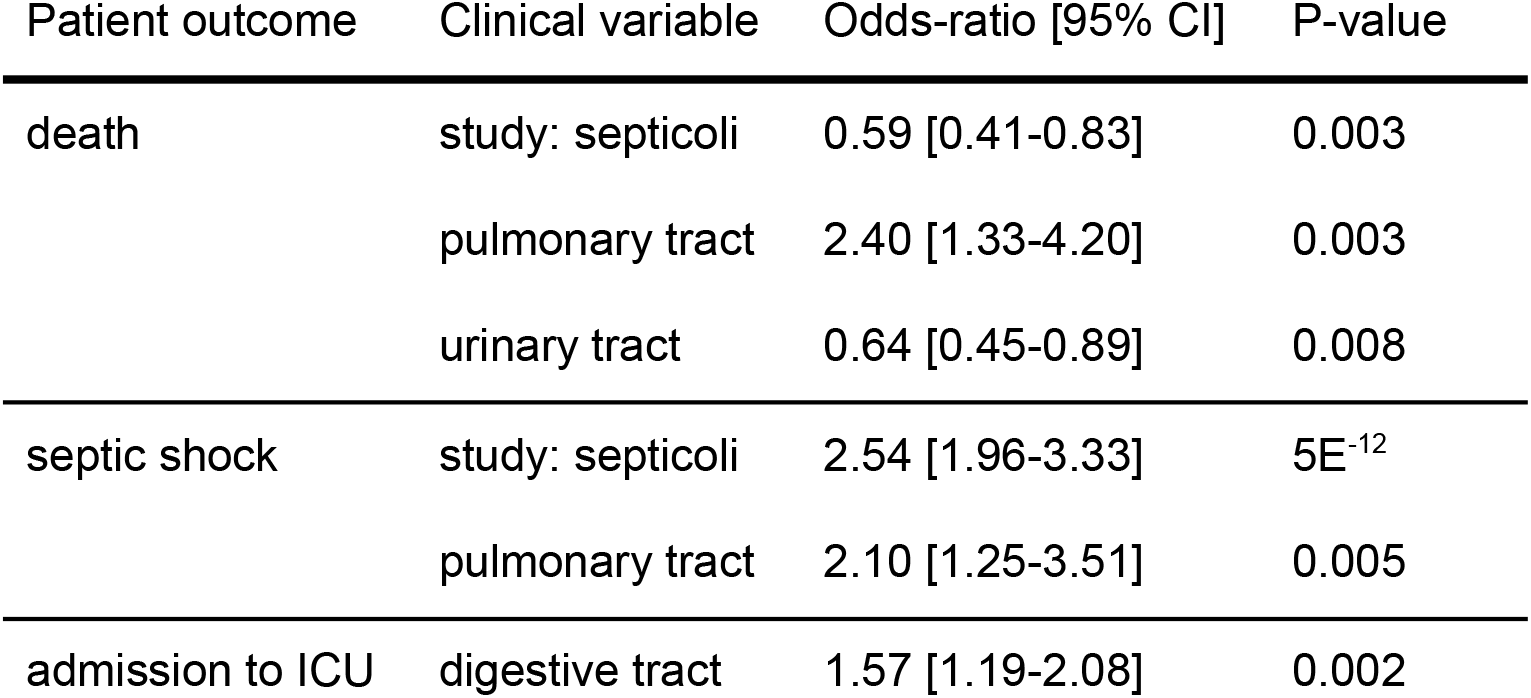
Multivariate analysis on the combined dataset. Only clinical variables with p-value < 0.01 are reported for each patient outcome, with the exception of the intercept. CI, confidence interval.

| Patient outcome | Clinical variable | Odds-ratio [95% CI] | P-value |
| --- | --- | --- | --- |
| death | study: septicoli | 0.59 [0.41-0.83] | 0.003 |
|  | pulmonary tract | 2.40 [1.33-4.20] | 0.003 |
|  | urinary tract | 0.64 [0.45-0.89] | 0.008 |
| septic shock | study: septicoli | 2.54 [1.96-3.33] | 5E<sup>-12</sup> |
|  | pulmonary tract | 2.10 [1.25-3.51] | 0.005 |
| admission to ICU | digestive tract | 1.57 [1.19-2.08] | 0.002 |

**Figure 2.**
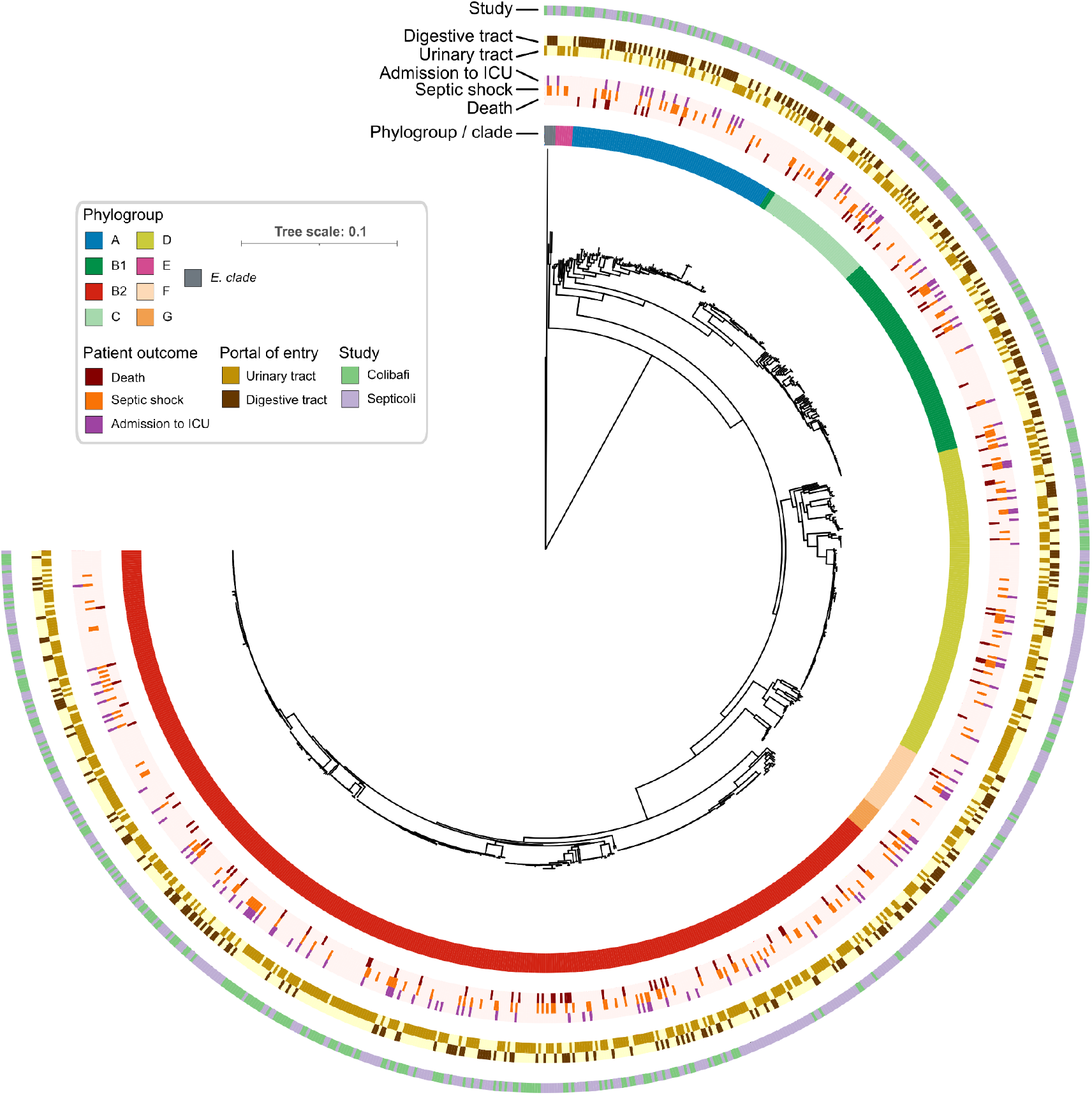
Core genome phylogenetic tree of the 910 *E. coli* isolates used in this study. Each ring reports the main bacterial and clinical variables of this study. The light color in the rings related to patient outcomes and portals of entry indicates the absence of the phenotype.

### Bacterial genetic factors can explain a significant fraction of the variation of the route of infection

We used narrow-sense heritability - the fraction of phenotypic variance that is explained by additive genetic effects^21^ - to estimate whether we could expect to find bacterial genetic variants in association with either patient outcome or the two main portals of entry. Since we found that clinical variables and the pathogen’s phylogroup are associated with our target variables, we measured heritability in three ways: using the phylogroup alone as a genetic effect^16^, and using a kinship matrix generated from the core genome phylogeny, alone or conditioning the analysis with the clinical variables in order to account for confounding factors (Figure 3). We found that phylogroups could explain 9% and 10% of the variation for the urinary and digestive tract infections, respectively (95% CI 0.5% - 46.6% and 0.6% - 46.7%, respectively), but none for any of the three patient outcomes. Variation in core genome SNPs could however explain 19% (95% CI 3.2% - 69.8%) of the variation in admission to the ICU, which we found to be negligibly reduced to 18% (95% CI 0% - 70.4%) when considering clinical covariates. While this may seem to indicate that the pathogen genetic variation might influence whether a patient will eventually need intensive care, we noted that this relatively high heritability was present in the Septicoli cohort alone (Figure 3b and 3c). We didn’t however find such a discrepancy between the two studies when we estimated the heritability for the portals of entry using either core SNPs alone or after conditioning; this indicates that there may be confounding factors that contribute to the decision to change a patient’s treatment which vary between the two studies. This is unsurprising, as the decision to admit a patient to the ICU can depend on the subjective assessment of a physician considering a patient’s comorbidities, as well as other subtle differences in care protocols. Conversely, the estimated heritability due to genetic effects for the portals of entry varies in magnitude between the combined dataset and the two cohorts alone, but we nonetheless found it to be > 0 in the three datasets. In particular, we found that genetic effects could explain 23% of the variance of both urinary and digestive tract portals of entry (95% CI 3.8% - 71.3% and 3.9% - 71.4%, respectively), which is more than double the variance explained by the isolates’ phylogroup (Figure 3a); this suggests that a genome-wide association analysis is likely to discover genetic variants associated with the portal of entry for BSI. This relatively high fraction of the phenotypic variability explained by genetic effects is however reduced when conditioning it on other clinical variables (10% and 11% for the urinary and digestive tract portal of entry, respectively, 95% CI 0% - 68.2% for both traits), which again underscores the influence of host characteristics in determining the establishment of bloodstream infections.

**Figure 3.**
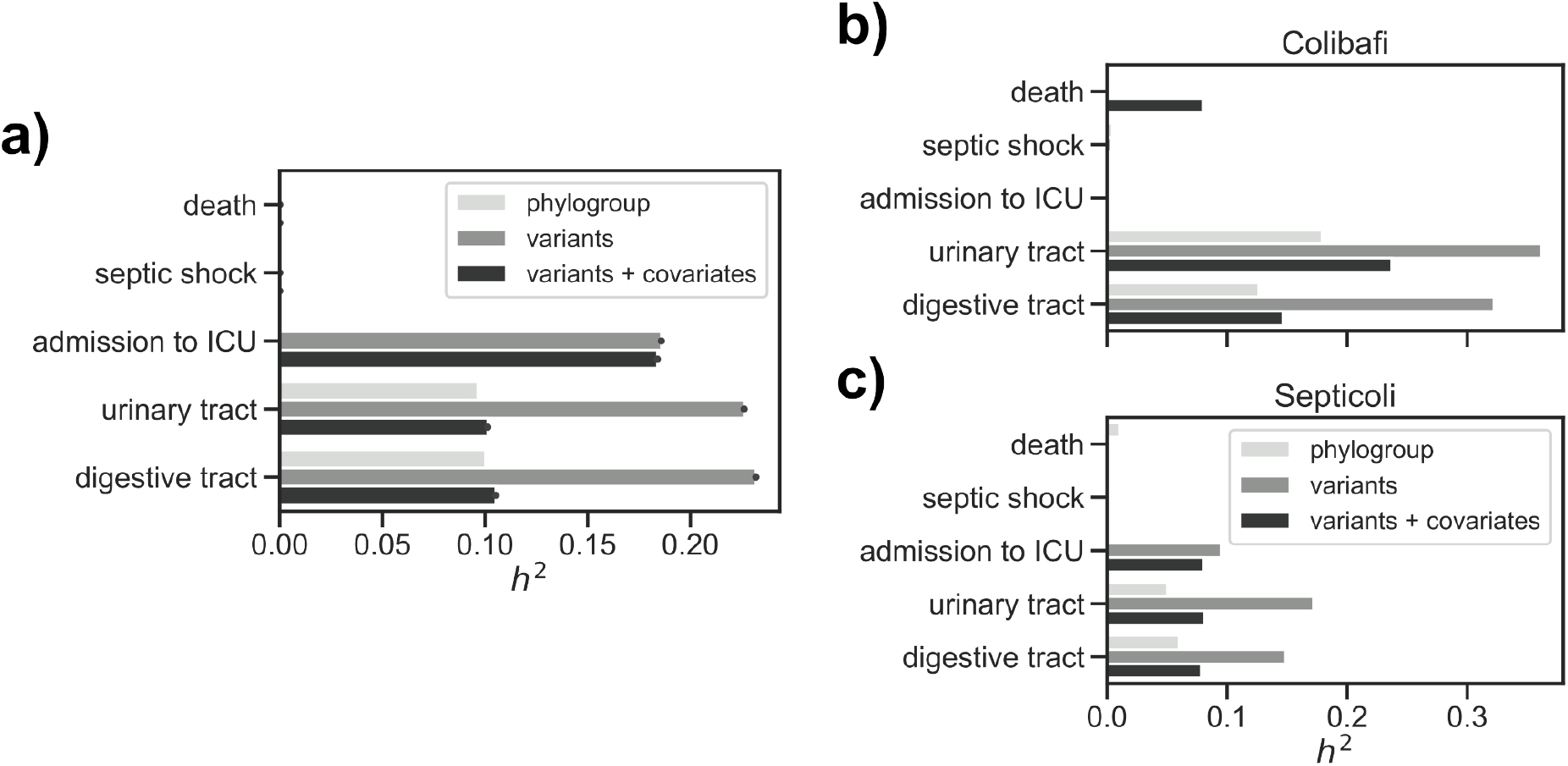
Narrow-sense heritability (*h*^*2*^) estimation for the target variables on the combined dataset. a) Heritability estimates in the two studies combined, using a covariance matrix generated from the isolates’ phylogroup (phylogroup), a kinship matrix generated from the core genome phylogeny (variants), and the same kinship matrix conditioned with the clinical variables (variants + covariates). b) Heritability estimates in the Colibafi and c) Septicoli cohorts alone.

### The *papGII/papGIII* operon is associated with the pathogen portal of entry

In order to account for both core and accessory genome genetic variability, which is one of the main differences between GWAS studies in human and bacterial datasets, we associated unitigs generated from a de Bruijn graph of all the bacterial isolates against the target variables^22,23^; namely the three patient outcomes and the two major portal of entry for BSI. We used a linear mixed model for the association, which has been shown to better correct for the influence of bacterial population structure in the association^24^. In order to account for the host and clinical factors on target variables, we conducted the association with the clinical variables as covariates^25^. Since our earlier analysis indicated that the portal of entry can influence patient outcome, we added this information as covariates when looking for bacterial genetic factors associated with the three patient outcomes.

Consistent with the heritability estimates, we found few or no unitigs associated after multiple testing correction with either the death of the patient (none for both the naÏve and conditioned association), the presence of septic shock (2 for the naÏve association and none for the conditioned association) and admission to ICU (1 for both the naÏve and conditioned association). Conversely, we found a large number of unitigs to be associated with either portal of entry; 975 and 1,061 for the urinary and digestive tract infections, respectively, when running a naÏve association, and a slightly lower number when adding clinical covariates, with 593 and 498 unitigs passing the significance threshold for the urinary and digestive tract, respectively (Figure 4a, Supplementary Table 3). Finding an association between individual unitigs and a phenotype of interest may be due to chance, even after multiple testing correction and the inclusion of covariates^26^; to reduce the influence of these factors on the results of the associations, we conducted a stringent analysis when mapping the unitigs back to each bacterial isolate; briefly, we took steps to exclude those unitigs that are mapped to multiple genes across all strains or that are found in a low number of strains (see Methods). After this stringent mapping step, we found no genes with associated unitigs mapped to them for the three patient outcomes, and 32 and 49 genes for the urinary and digestive tract portal of entry, respectively and independently on whether we used the clinical covariates in the unitig association step (Figure 4b, Supplementary Table 4 and Supplementary Material 1). The absence of any associated gene with the three patients’ outcomes is in agreement with the heritability estimates, and with our argument that the relatively high heritability for the admission to the ICU may be the result of confounders.

**Figure 4.**
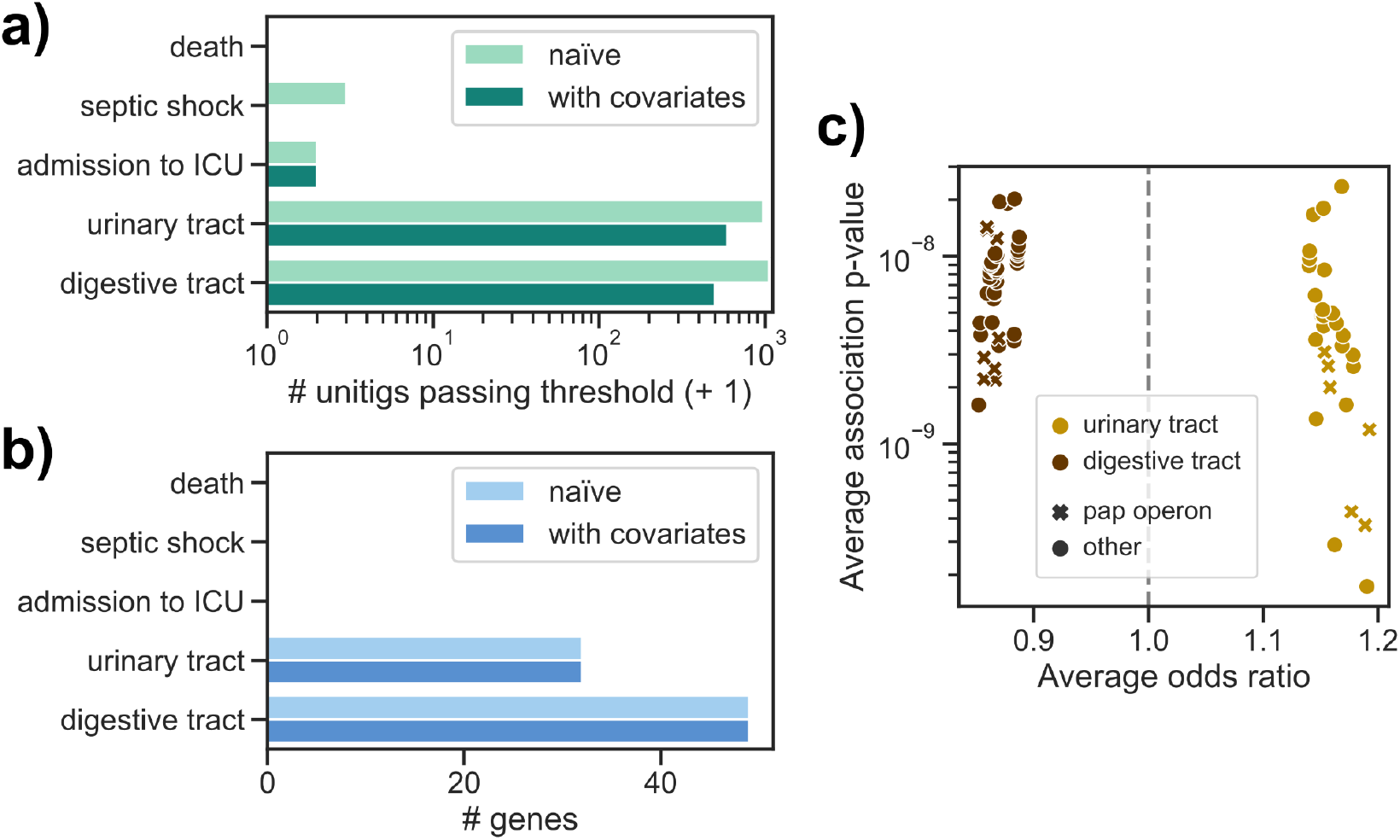
Genome-wide association analysis results on the combined dataset. a) Number of unitigs passing the multiple testing correction p-value threshold for each target phenotype. b) Number of genes with significantly associated unitigs mapped to them for each target phenotype. c) Average odds-ratio and association p-value for each gene in the two portals of entry.

We found a large overlap in associated genes for the urinary and digestive tract portals of entry, with 25 genes shared between them. Furthermore, we observed similar effect sizes reported for those genes in the two phenotypes (Pearson’s r for the average odds-ratio 0.75), but with opposite signs, which is likely the result of the two phenotypes being almost exactly mutually exclusive (Figure 4c). Among these 25 genes, we found that 10 belonged to the *pap* operon or in its immediate vicinity; the genes in this operon encode for a type P pilus, which has been shown to interact with glycolipids present on uroepithelial cells and is therefore believed to be one of the main defining loci for severe UTI. We found that the *papGII* and *papGIII* variants of the *papG* gene encoding for the adhesin part of the tip were associated with both portals of entry (Supplementary Figure 1). The PapGII adhesin is mainly found in acute pyelonephritis and binds preferentially to Gb4 (GalNAcβ1-3Galα1-4Galβ1-4GlcCer), which is abundant in the upper urinary tract of humans. P-pili presenting PapGIII are common in human cystitis, but rare in pyelonephritic isolates; they bind Gb5 (GalNAcα1-3-GalNAc3Galα1-4Galβ1-4GlcCer)^27^. We found another 4 genes associated with both portals of entry and encoded in the vicinity of the *pap* operon, all with high sequence similarity (blastp sequence identity > 95%) to genes annotated as phosphoethanolamine transferases, or *opgE*. This gene is involved in the biosynthesis of osmoregulated periplasmic glucans (OPGs), which in turn regulate motility and secretion of exopolysaccharides and are considered virulence factors for Gram-negative species^28–31^. We found these putative *opgE* genes encoded in the vicinity of phage-derived integrase genes, which are also associated with the portals of entry (annotated as *intA* and *intS*). We found that the putative *opgE* gene was encoded in the near vicinity of the *pap* operon (distance < 15kbp) in 118 strains, and an even shorter distance (< 10kbp) between the *pap* operon and the edge of its contig for those strains (213) in which the *pap* operon and the putative *opgE* gene were encoded in separate contigs (Supplementary Figure 1). We therefore concluded that both the putative *opgE* gene and the integrase genes are part of the same genetic island that may have been acquired through horizontal gene transfer across *E. coli* strains^13^.

### A larger sample size could reveal additional bacterial factors involved in bacteremia

Our heritability estimates and association results are in good agreement both with previous results about the difficulty of finding bacterial genetic elements associated with virulence from clinical cohorts^16^ and with the importance of the *pap* operon in enabling severe UTI^13^. We next asked whether it would theoretically be possible to find even more associations from cohorts measuring *E. coli* BSI; would an increase in sample size lead to the discovery of more bacterial genetic factors able to affect the establishment and the outcome of BSI? To answer this question, we generated a dataset of 10,000 simulated genomes - one order of magnitude higher than the dataset presented in this study - with mutation and recombination rates similar to those of *E. coli*, and two phenotypes with either “high” or “low” heritability (0.2 and 0.05, respectively)^26^. For each phenotype we selected 28 causal variants with a range of effect sizes. We then ran a GWAS on the full dataset and in two smaller samples, in order to determine the empirical statistical power (Figure 5). We found that in this simulated dataset an increase in sample size by an order of magnitude would be needed to discover most of the causal variants (mean recall 57%) for the phenotype with high heritability, which is a large increase from the sample size most similar to this study (1,000 samples, mean recall 5%). Conversely, we found that for the low heritability scenario only a relatively low statistical power (mean recall 10%) could be achieved with a large sample size of 10,000 samples, and no power when using 1,000 samples (mean recall 0). While this simulation cannot be directly compared with the genetics of complex bacterial phenotypes such as BSI caused by *E. coli*, it points to the theoretical possibility of further refining these results if a larger set of samples could be assembled. This could prove particularly fruitful if patient outcomes are indeed influenced at least partially by bacterial genetic factors.

**Figure 5.**
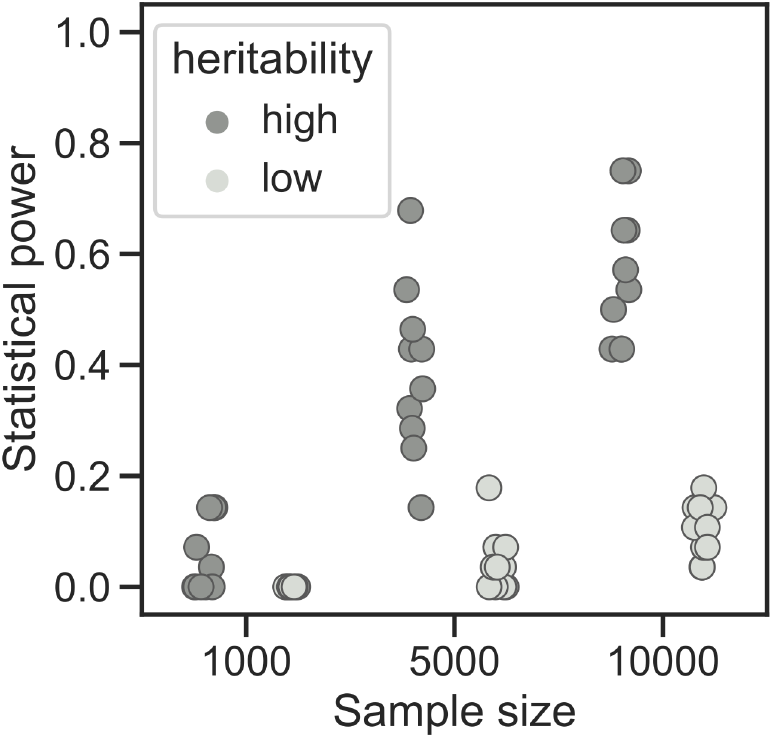
Power simulations. The proportion of causal variants passing the significance threshold is reported for each sample size and heritability for the simulated phenotypes.

## Discussion

In this study we leveraged the clinical and genetic data of two very similar BSI clinical cohorts in order to test whether *E. coli*’s genetic variation has an influence on the course of severe bloodstream infections. As opposed to our previous analysis using a well-controlled mouse model of sepsis where GWAS identified iron capture systems as main drivers of virulence^20^, we did not find a clear locus effect for the three patient outcomes tested here. This is in agreement with a previous work in which we used 60 *E. coli* strains derived from bacteraemic patients and tested their virulence in the mouse model, looking for genetic determinants for the clinical severity of infection. Indeed, virulence based on an animal model was correlated with bacterial virulence determinants but not with pejorative clinical outcome of BSI^32^. In fact the animal model is a controlled environment, as the individual tested are healthy and homogeneous (same sex, age, weight and diet); furthermore the standardized inoculation uniformizes the portal of entry, thus allowing for an unbiased evaluation of the intrinsic virulence of each strain^33^. The data presented here once more seems to point to either a negligible influence of bacterial genetic variation on infection outcomes when compared to host and clinical factors, a complex trait influenced by multiple loci, or to a lack of statistical power due to a relatively low sample size.

The results from the heritability analysis from this dataset of combined cohorts is mixed in this regard, as we found that the variance in a patient’s death or septic shock is not explained by bacterial genetics, while we found that locus effects may explain up to 19% of the variance in admission to ICU. When we broke down this analysis in the two cohorts alone, we observed that this relatively high heritability is only observed in the Septicoli cohort; as the decision to change the care of a patient is a complex decision dependent on the subjective assessment of clinicians and other hospital-specific policies, we believe that this high estimate may be the result of confounders. A more objective measure of disease burden may therefore be needed in order to properly test for the influence of bacterial genetics on BSI outcomes, together with an increase in sample size, as suggested by our simulations.

On the other hand, we found a clear association between the *pap* operon and surrounding genes and the route of entry for BSI. This agrees with an earlier study with a similar sample size that looked specifically at invasive UTI^13^. We can point to a common theme in the genome-wide association studies so far conducted in *E. coli* infection models: the main associated genetic elements come from fairly frequent (∼50% of the population) pathogenicity islands that have been previously described, sometimes decades before the ubiquity of genomic data made GWAS studies feasible^34–39^. One can then wonder whether these approaches are likely to ever lead to the discovery of previously undescribed genetic variants able to modulate the establishment of disease and its outcomes. We argue that as genomic sequencing of pathogens is becoming a routine part of clinical or epidemiological practice^40–42^, we will likely eventually reach very large sample sizes, similar to what is currently available for human GWAS studies^43^, and possibly larger, as has been recently shown for Sars-Cov-2 genomic epidemiology efforts^44,45^. Apart from increasing the power to discover genetic variants associated with a phenotype, a large sample size would allow for the discovery of rare or ultra-rare variants, which in turn may have a relatively large influence on the phenotype of interest, alone or collectively^23^, as has recently been appreciated in the study of human traits and disease^46^. In the context of bacterial infection, in which we and others have shown how host factors contribute to a large extent, a further help will likely come from including the host genetic variation into the association; a joint human/bacterial association analysis may however require an even larger sample size in order to account for potential interactions between host and bacterial genetic elements^16^. Taken together, the assumed inevitability of clinical genome sequencing together with careful recording of host and clinical data may eventually lead to comprehensively cataloging the fraction of *E. coli* genetic variants that influence bloodstream infections.

## Materials and methods

### Dataset

The Colibafi and Septicoli studies were prospective observational cohort studies conducted in tertiary-care teaching hospitals in the Paris area. Adult patients with *E. coli* BSI were included. Only patients previously included in the study for a previous BSI episode, and patients receiving vasopressors before the onset of BSI were excluded. *E. coli* BSI was defined as the isolation of *E. coli* from at least 1 blood culture bottle. Data were prospectively collected by clinicians in each centre on two separate visits: Visit 1 corresponded to the time of BSI (the day the blood culture was drawn; data were collected retrospectively 24-48h hours later, once the blood culture had grown) and Visit 2 corresponded to the day of discharge or in-hospital death (or day 28 if the patient was still hospitalized). For each episode, the first *E. coli* strain collected in the blood culture was identified. The primary endpoint was vital status at discharge or Day-28 (i.e. Visit 2). In each centre, an infectious diseases clinician and a microbiologist were in charge of including patients and completing the case report form (see Colibafi and Septicoli groups in the Acknowledgments section). A steering committee was in charge of implementation and a scientific committee responsible for scientific overview.

From the combined dataset we removed those variables with more than 15% of missing values (whether the patient had received a transplant, neutropenia, pregnancy status, body mass index, patient discharge route), and we added a binary variable to record the study provenance of each sample. We imputed the remaining missing values using the MICE package, v3.12.0, using 15 iterations^47^. The raw and imputed combined datasets’ summary statistics are available as Supplementary Table 1.

### Univariate and multivariate analysis

We tested the association between clinical variables and patient outcome in a similar way as it was done in the original studies^8,10^; briefly, we first applied a min-max scaler to the age variable to bring it in the [0-1] range. For each patient outcome we then tested each clinical variable using a logistic regression as implemented in the statsmodels package, v0.11.1, using the study provenance as a covariate. We used those variables with association p-value < 0.1 to run a multivariate logistic regression, using a backward stepwise selection method to construct the final model, using the MASS package v7.3_51.3^48^.

### Whole genome sequencing and annotation

Bacterial genomes were sequenced using Illumina NextSeq technology as previously described^19^. The genomes from the Colibafi and Septicoli collections are available (Bioproject PRJEB39260 and PRJEB35745, respectively). All genomes were assembled with shovill version 1.0.4 using SPAdes v3.13.1^49^ and standard parameters, and then annotated with Prokka 1.14.5^50^. A phylogenetic tree was computed from a core genome multiple sequence alignment, as computed by Roary v3.12^51^, using IQ-TREE v1.6.12^52^, under the GTR+F+I+G4 model. The tree was visualized using the iTOL web interface^53^. We collapsed all genes encoded in the sequenced genomes into gene families using panaroo v1.2.4^54^ with default parameters.

### Heritability estimates

We estimated narrow-sense heritability for the five target variables, using 2 different covariance matrices; one built from the phylogroup membership of each strain and another using a kinship matrix built from the core genome phylogeny, in which the distance from the most recent common ancestor is used for each pair of samples. For the latter covariance matrix we also used the same clinical covariates as in the GWAS analysis (see below). We used Limix v3.04^55^, assuming normal errors for the point estimate and we computed the 95% confidence intervals using the ALBI package (commit 90d819e)^56^.

### Association analyses

We derived unitigs by constructing a compressed de Bruijn graph from the input genomes, using unitig-counter v1.1.0^22,23^. We computed the distance between each pair of samples by using mash 2.2.2^57^ with a sketch size of 10,000; we used the resulting distance square matrix to compute associations between phylogroups and each target variable, using pyseer v1.3.6^58^. We tested for locus effects using the unitigs presence/absence vector with the FastLMM^59^ linear mixed-model and a kinship matrix derived from the core genome phylogeny, using pyseer v1.3.6^58^. We run two associations; a “naÏve” one that accounted for population structure only, and one additionally conditioning on the clinical variables (“with covariates”). For the three patient outcomes we used all available variables as covariates with the exception of “death”, “septic shock” and “admission to ICU”, but including the portals of entry, which were excluded when those were the target variables. All the clinical variables used as covariates are described in Supplementary Table 1. We determined a significance threshold by counting the number of unique unitigs presence/absence patterns tested, which reduces the risk of excessively deflating association p-values. We mapped the unitigs passing the significance threshold back to all input genomes and their genes using bwa v0.7.17-r1188^60^ and bedtools v2.30.0^61,62^, using the output of panaroo to assign each unitig to a gene cluster. The unitigs were further filtered to reduce the number of spurious associations: unitigs were excluded if they were shorter than 30bp, if they were mapped to multiple locations in each genome, if they mapped to less than 9 samples (∼1% of the total sample size) and if they were mapped to more than 10 different genes across all samples. We further annotated the gene families with mapped unitigs by taking a representative protein sequence from all genomes encoding each gene and using it as an input for eggnog-mapper v2.1.3^63^.

We tested for the association of rare variants (minimum allele frequency < 1%) by performing a burden test, that is, we performed associations between deleterious rare variants in each gene separately and the five target phenotypes. We derived short variants from each sample against the complete genome of *Escherichia coli* IAI39 - which belongs to phylogroup B2 - using snippy v4.6.0 and annotated them using SnpEff v5.0^64^. We then merged the individual VCF files and filtered for rare variants using bcftools v1.13^65^. We further filtered the resulting variants according to their annotation: variants annotated as “disruptive”, “frameshift”, “start codon loss”, “stop codon gain”, and “stop codon loss”; for missense variants we assessed the likelihood that they were deleterious to protein function using the SIFT algorithm, as implemented in the SIFT4G package v2.0.0^66^, using the uniref50 subset of the Uniprot database^67^ (downloaded on June 16, 2021) to construct the multiple sequence alignments. We considered a missense variant to be deleterious if the protein residue had a median information content below 3.25 and score < 0.05. The association was run in a similar way as the one with common unitigs (linear mixed model and clinical covariates) using pyseer v1.3.6^58^. No significant hit was found with this association method.

### Power simulations

We performed a statistical power analysis to test whether an increase in sample size could lead to the discovery of additional variants associated with a binary phenotype with heritability similar to that estimated in this study. We used the BacGWASim package v2.1.1^26^ to generate both simulated variants and phenotypes. We simulated 10,000 bacterial genomes each 1,000,000 bp long, using a mutation rate of 0.06 and recombination rate of 0.01. We then simulated two binary phenotypes: one with a “high” (0.2) and one with a “low” (0.05) heritability; for both phenotypes we assumed a prevalence of 50% and generated 10 sets of 28 causal variants with minimum allele frequency of 10%. For each batch of simulated phenotypes we ran an association with pyseer v1.3.6^58^ using logistic regression and population structure correction using the first four components of the multidimensional scaling obtained from the samples pairwise distance matrix computed using mash v2.2.2^57^. Statistical power was computed as the proportion of causal variants that passed the significance threshold, computed by counting the number of unique presence/absence patterns for all tested variants.

## Data Availability

All data produced in the present work are contained in the manuscript and are available in public repositories such as ENA and github

https://github.com/microbial-pangenomes-lab/2021_ecoli_pathogenicity

## Code availability

Apart from the software packages mentioned in the previous sections, the following were used to run the analysis and generate the visualizations presented in this work: pandas v1.2.2^68^, numpy v1.20.0^69^, scipy v1.6.0^70^, matplotlib v3.3.4^71^, seaborn v0.11.1^72^, biopython v1.79^73^, reportlab v3.5.68^74^, gffutils v0.10.1, jupyterlab v3.0.7^75^. Most of the analysis were incorporated in a reproducible pipeline using snakemake v6.5.0^76^ and conda v4.10.3^77,78^, which is available as a code repository on GitHub (https://github.com/microbial-pangenomes-lab/2021_ecoli_pathogenicity) under a permissive license (MIT).

## Ethics statements

Both multicenter clinical trials were approved by ethic committees. The Colibafi study was approved by the French Comité de Protection des Personnes of Hôpital Saint-Louis, Paris, France (approval #2004-06, June 2004). The Septicoli study was approved by the French Comité de Protection des Personnes Ile de France n°IV (IRB 00003835, March 2016) and was registered on clinical trials in September 2016 (ClinicalTrials.gov Identifier: NCT02890901). Because of their non-interventional nature, only an oral consent from patients was requested under French law. Both studies conformed to the principles of the Helsinki declaration.

## Acknowledgements

We thank John Lees and Lars Barquist for providing suggestions on the final version of the manuscript. We are grateful to Jean-Marie Lacroix for having kindly shared his knowledge on opgE. This work was partially funded by a Translational Research Grant from the Agence Nationale de la Recherche (ANR), Ministry of Higher Education and Research, France 2015 (grant n°ANR-15-CE-17-0019-01). ED was partially supported by the “Fondation pour la Recherche Médicale” (Equipe FRM 2016, grant number DEQ20161136698). GR was supported by a “Poste d’accueil” funded by the “Assistance Publique-Hôpitaux de Paris” (AP-HP) and the “Commissariat à l’énergie atomique et aux énergies alternatives” (CEA) personal grant for his PhD. MG was supported by the Deutsche Forschungsgemeinschaft (DFG, German Research Foundation) under Germany’s Excellence Strategy - EXC 2155 - project number 390874280.

The Colibafi Group is composed of: Michel Wolff, Loubna Alavoine, Xavier Duval, David Skurnik, Paul-Louis Woerther, Antoine Andremont, Etienne Carbonnelle, Olivier Lortholary, Xavier Nassif, Sophie Abgrall, Françoise Jaureguy, Bertrand Picard, Véronique Houdouin, Yannick Aujard, Stéphane Bonacorsi, Agnès Meybeck, Guilène Barnaud, Catherine Branger, Agnès Lefort, Bruno Fantin, Claire Bellier, Frédéric Bert, Marie-Hélène Nicolas-Chanoine, Bernard Page, Julie Cremniter, Jean-Louis Gaillard, Françoise Leturdu, Jean-Pierre Sollet, Gaëtan Plantefève, Xavière Panhard, France Mentré, Estelle Marcault, Florence Tubach, Olivier Clermont.

The Septicoli Group is composed of: Virginie Zarrouk, Frederic Bert, Marion Duprilot, Véronique Leflon-Guibout, Naouale Maataoui, Laurence Armand, Liem Luong Nguyen, Rocco Collarino, Anne-Lise Munier, Hervé Jacquier, Emmanuel Lecorché, Laetitia Coutte, Camille Gomart, Ousser Ahmed Fateh, Luce Landraud, Jonathan Messika, Elisabeth Aslangul, Magdalena Gerin, Alexandre Bleibtreu, Mathilde Lescat, Violaine Walewski, Frederic MechaÏ, Marion Dollat, Anne-Claire Maherault, Michel Wolff, Mélanie Mercier-Darty, Bernadette Basse, Olivier Clermont.

## Author’s contributions

MG, ED, AL and VL conceived the study. MEF, AL and VL provided support for the analysis of the clinical variables. GR and OC assembled and annotated the bacterial genomes and generated the core genome phylogenetic tree. MEF and CL provided support for the univariate and multivariate analysis. MG prepared the data for all association analysis and ran them. BC annotated the associated genes. MG generated all visualizations and tables. MG and ED wrote the manuscript. All authors read and approved the current version of the manuscript.

## Supplementary figures

**Supplementary Figure 1.**
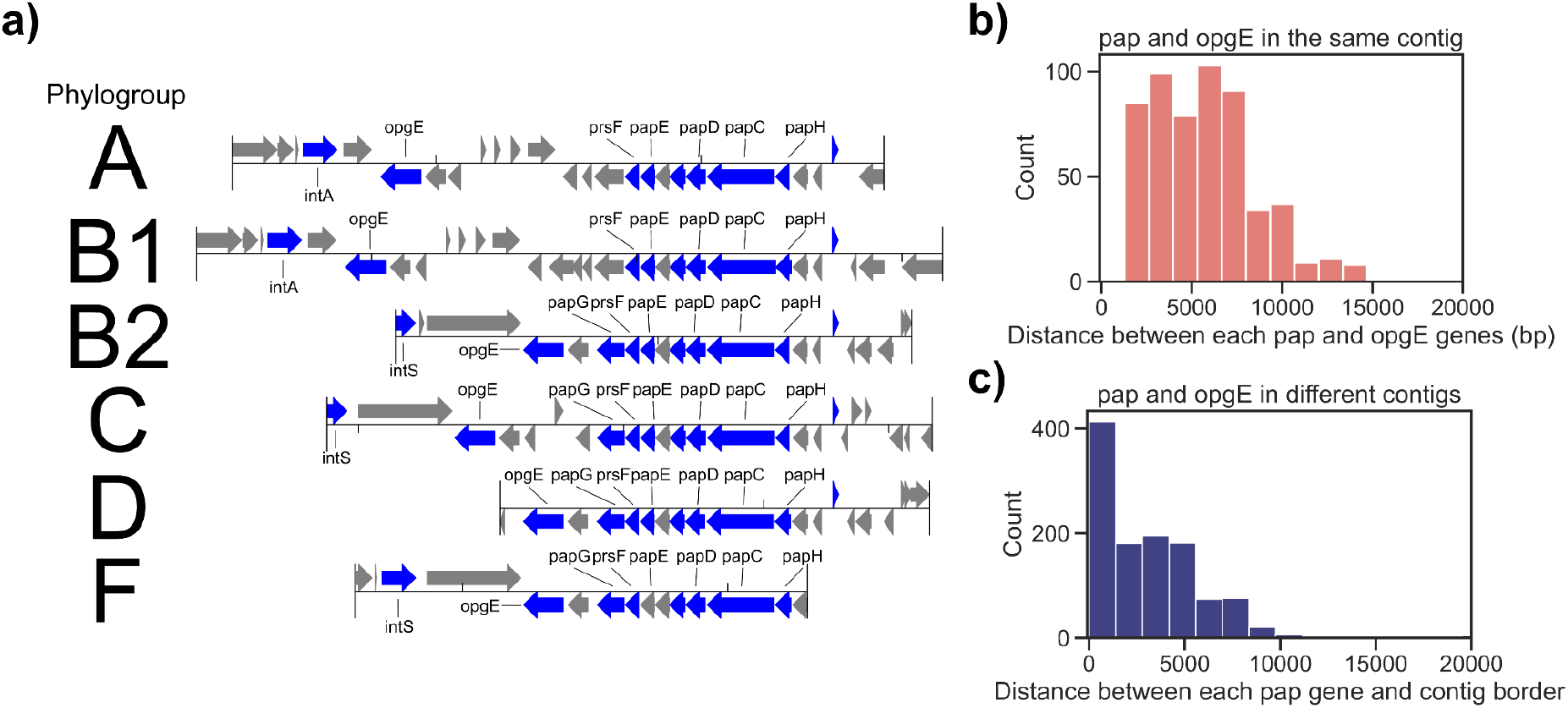
Structure of the *pap* operon island and relative position of the putative *opgE* gene. a) Position and relative orientation of the *pap* operon and the putative *opgE* gene is shown for one sample strain belonging to each major *E. coli* phylogroup. Genes colored in blue have at least one associated unitig mapped to it (using the entry through the urinary tract as target variable), grey otherwise. b) Distance between the putative *opgE* gene and the *pap* operon in those strains in which the two genetic elements are encoded in the same contig, and c) Distance between the *pap* operon and the edge of the contig in those strains in which the putative *opgE* gene is encoded in a different contig.

## Supplementary materials

- Supplementary Table 1: clinical variables for both cohorts, in its original form and after imputation of missing values
- Supplementary Table 2: lineage associations
- Supplementary Table 3: associated unitigs
- Supplementary Table 4: genes to which associated unitigs map to (see Methods for mapping and filtering)
- Supplementary Material 1: aminoacid sequence for each associated gene, sampled randomly for each gene cluster

## Notes

### Competing Interest Statement

The authors have declared no competing interest.

### Author Declarations

Both multicenter clinical trials were approved by ethic committees. The Colibafi study was approved by the French Comite' de Protection des Personnes of Hopital Saint-Louis, Paris, France (approval #2004-06, June 2004). The Septicoli study was approved by the French Comite' de Protection des Personnes Ile de France n IV (IRB 00003835, March 2016) and was registered on clinical trials in September 2016 (ClinicalTrials.gov Identifier: NCT02890901). Because of their non-interventional nature, only an oral consent from patients was requested under French law. Both studies conformed to the principles of the Helsinki declaration.

